# Determinants of Success in Hyperbaric Oxygen Combination Therapy for Sudden Hearing Loss

**DOI:** 10.1101/2025.08.01.25332735

**Authors:** Jie Chen, Jiaoni Gong, Wangwang Hu, Beier Tong, Jiabin Huang, Lu Shi

## Abstract

**Objective:** Hyperbaric oxygen (HBO) therapy plays a crucial role in the treatment of sudden hearing loss (SHL). This study aims to discuss various factors influencing treatment outcomes and examine the treatment protocol of HBO combination therapy for SHL.

**Methods:** From May 2023 to April 2024, 87 SHL patients were categorized into 4 groups: the Cure Group (CG), Significant Effect Group (SG), Improved Group (IMG), and Invalid Group (ING). We conducted pairwise multiple comparisons and ordered logistic regression analysis to identify factors with statistically significant associations and to evaluate their relationship with clinical outcomes.

**Results:** By pairwise multiple comparisons, CG patients were younger than IMG and ING(P < 0.05). SG had a longer treatment course than IMG and ING(P < 0.05). ING had the longest time from onset to treatment(P < 0.05). SG and IMG were more likely to dizzy than CG(P < 0.05).As for the types of audiometric curves, the total effective rate of low frequency descent type was higher than any other types(P < 0.05). As the results of ordered logistic regression analysis shown, low frequency descent type and appropriate long treatment course were protective factors to prognosis(P < 0.05). While the elder patients and the longer time from onset to treatment were risk factors(P < 0.05).

**Conclusion:** The factors such as low frequency descent type,younger age,an appropriately longer treatment duration, absence of dizziness and timely initiation of treatment were relevant to a good clinical outcome. A treatment course of 10 to 20 HBO therapy might be more suitable.

## Introduction

Sudden hearing loss (SHL) is considered to be an otologicemergency, usually accompanied by tinnitus and/or dizziness. Microcirculation disturbance, viral infection, autoimmunity, and stress are some of the suggested theoretical causes.^[1-3]^ But most SHL is defined as no identifiable cause. Much of the literature indicates that 32-65% of SHL patients may recover spontaneously.^[4, 5]^ However, cinical experience shows that these numbers may be overestimated. If not recognized and treated promptly, it may result in persistent hearing loss and tinnitus.^[6]^ Whlie the direct mortality rate of SHL is very low, the impact on quality of life is significant. That is, the most effective and the safest therapies for the patients is crucial for improving prognosis and minimizing the risk of complications.^[7]^ Currently, commonly used medications in clinical practice include glucocorticoids, neurotrophic agents, vasodilators, anticoagulants, antioxidants, and others.

Hyperbaric oxygen (HBO) therapy is defined as the process of patients inhaling 100% pure oxygen at an environment above 1 atmosphere(ATA) to treat disease.^[8, 9]^ HBO was first used in the treatment of SHL in the 1960s, but failed to show advantage due to sample selection bias.^[10-12]^ With the continuous deepening of HBO research, in the past 20 years, there have been important breakthroughs in some concepts and mechanisms of HBO. The mechanisms of HBO treatment can effectively address most of the pathogenesis of SHL. In recent years, numerous clinical studies have demonstrated that HBO combination therapy is significantly more effective than drug therapy alone. ^[13]^ However, there was currently limited and incomplete research on the prognostic factors of HBO combination therapy. Whether a longer treatment course of treatment would lead to better outcomes was also worth considering. Our paper presents a retrospective analysis of clinical data from 87 SHL patients who received HBO combination therapy to discuss various factors influencing treatment outcomes and examine the treatment protocol of HBO combination therapy for SHL.

## Methods

### Study Population

From 1 May 2023 to 30 April 2024, a retrospective cohort study was conducted involving 87 consecutive patients diagnosed with SHL who underwent HBO therapy at our hospital. Participants were anonymously enrolled through a standardized screening process of electronic medical records, with all identifiers removed prior to analysis. All patients were satisfied relevant diagnostic criteria for SHL. ^[5]^ Inclusion criteria:. (1) the first occurred, (2) seeking medical attention at our hospital within 72 hours of onset and receiving standardized treatment (as specified in the “Therapy” section below), (3) age > 18 years, (4) idiopathic neurosensory hearing loss with unilateral or bilateral involvement, (5)normal function of eustachian tube. Exclusion criteria: (1) previous otologic surgery in the ear, (2)previous history of cancer, (3)untreated hypertension and diabetes, (4)patients unable to cooperate with treatment.

Some baseline characteristics were collected as follows: age, sex, treatment course, the time from onset to treatment, hypertension, diabetes, hyperlipidemia, smoking, drinking, with dizziness/tinnitus or not, the types of audiometric curves, and the report of electrical audiometry before and after treatment. Based on the degree of hearing impairment assessed using the average hearing level at frequencies of 250, 500, 1000, 2000 and 4000 Hz during the initial visit, we divided the patients into 4 groups (30-50dB, 51-70dB, 71-90dB, >90dB).

### Therapy

The patient received treatment with 100% oxygen at a pressure of 2 ATA for a total duration of 100 minutes, which included 15 minutes of air compression, 30 minutes of oxygen breathing, a 5-minute rest period, followed by another 30 minutes of oxygen breathing, and concluding with 20 minutes of air decompression. This treatment was administered once daily, with breaks on weekends.

Glucocorticoid Therapy:1. Initial phase: Methylprednisolone 80 mg intravenous infusion daily for 3 days; 2. Subsequent phase: Reduce to 40 mg intravenous infusion daily for an additional 3 days;3. Maintenance phase: Transition to oral administration with gradual dose tapering.

Batroxobin Injection Protocol:1. Initial dose: 10 BU intravenous infusion; 2.Subsequent dosing: 5 BU administered every other day, based on coagulation profile monitoring (prothrombin time and fibrinogen levels);3.Treatment course: Total of 5 doses constitutes one complete therapeutic cycle.

Additional therapeutic measures comprised adjunctive pharmacotherapy (including non-specific medications) and targeted management of pre-existing comorbidities.

### Treatment assessment

After treatment, a return of the average hearing level across the 5 frequencies (250, 500, 1000, 2000, and 4000 Hz) to pre-onset levels or normal was classified as a cure (CG). A hearing gain of more than 30 dB at the end of treatment was considered a significant effect (SG). Hearing gains between 15 and 30 dB were categorized as improvement (IMG), while gains of less than 15 dB were deemed ineffective (ING). The total effective rate included the first three categories (CG + SG + IMG).

### Statistics

Continuous variables were expressed as the mean value ± standard deviation or medians with interquartile ranges. Categorical variables were expressed as proportions. Baseline characteristics were analyzed with ANOVA, Kruskal-Wallis test and Chi-square test. Multiple samples between each other were compared by LSD-t test and Kruskal-Wallis test followed by all pairwise multiple comparisons. Variables with a P value <0.10 were selected for ordered logistic regression analysis. P value was considered significant <0.05. All statistical analyses were performed using SPSS version 26.0 (SPSS Inc, Chicago, IL).

## Results

Of the 87 patients screened, 18 were classified as cured (CG), 29 significant effective (SG), 24 improved (IMG), and 16 invalid (ING). By comparison, 4 groups were similar in sex, hypertension, diabetes, hyperlipidemia, smoking, drinking, with tinnitus or not. We observed statistical significance with a P value < 0.05 in age, treatment course, the time from onset to treatment, and with dizziness or not (Table 1).

**Table 1:** Comparison of groups.

| Characteristics | CG(*n* = 18) | SG(*n* = 29) | IMG(*n* = 24) | ING(*n* = 16) | *P* |
| --- | --- | --- | --- | --- | --- |
| Age(years),mean(SD) | 42.56(15.86) | 50.59(13.99) | 54.00(14.32) | 54.88(12.80) | 0.043* |
| Male(%) | 8(44.4) | 13(44.8) | 12(50.0) | 8(50.0) | 0.970 |
| course(times),median(IQR) | 7.5(4.8-10.3) | 10(7-20) | 7(4.25-8) | 5(3-7) | 0.001* |
| the time from onset to treatment(days),mean(SD) | 7.22(2.71) | 6.24(3.11) | 8.54(11.44) | 15.13(18.15) | 0.040* |
| Hypertension(%) | 3(16.7) | 3(10.3) | 8(33.3) | 6(37.5) | 0.087 |
| Diabetes(%) | 1(5.6) | 5(17.2) | 7(29.2) | 4(25.0) | 0.204 |
| Hyperlipidemia(%) | 4(22.2) | 14(48.3) | 11(45.8) | 9(56.3) | 0.196 |
| Smoking(%) | 3(16.7) | 3(10.3) | 6(25.0) | 4(25.0) | 0.467 |
| drinking(%) | 1(5.6) | 2(6.9) | 4(16.7) | 5(31.3) | 0.110 |
| comorbidity |  |  |  |  |  |
| tinnitus(%) | 15(83.3) | 24(82.8) | 22(91.7) | 11(68.8) | 0.326 |
| dizziness(%) | 2(11.1) | 15(51.7) | 14(58.3) | 4(25.0) | 0.005* |

CG:the Cure Group; SG:Significant Effect Group; IMG:Improved Group; ING:Invalid Group
\*, $P < 0.05$

By pairwise multiple comparisons of indicators above, CG were younger than IMG and ING(42.56±15.86 vs 54.00±14.32,P=0.012; 42.56±15.86 vs 54.00±14.32,P=0.014). SG had a longer treatment course than IMG and ING[10(7-20) vs 7(4.25-8),P=0.004; 10(7-20) vs 5(3-7),P<0.001]. But there were no statistical difference between CG and SG in age and treatment course. ING had the longest time from onset to treatment (7.22±2.71 vs 15.13±18.15,P=0.024; 6.24±3.11 vs 15.13±18.15,P=0.006; 8.54±11.44 vs 15.13±18.15,P=0.045). SG and IMG were more likely to dizzy than CG[15(51.7) vs 2(11.1),P=0.036; 14(58.3) vs 2(11.1),P=0.013](Table 2).

**Table 2:**
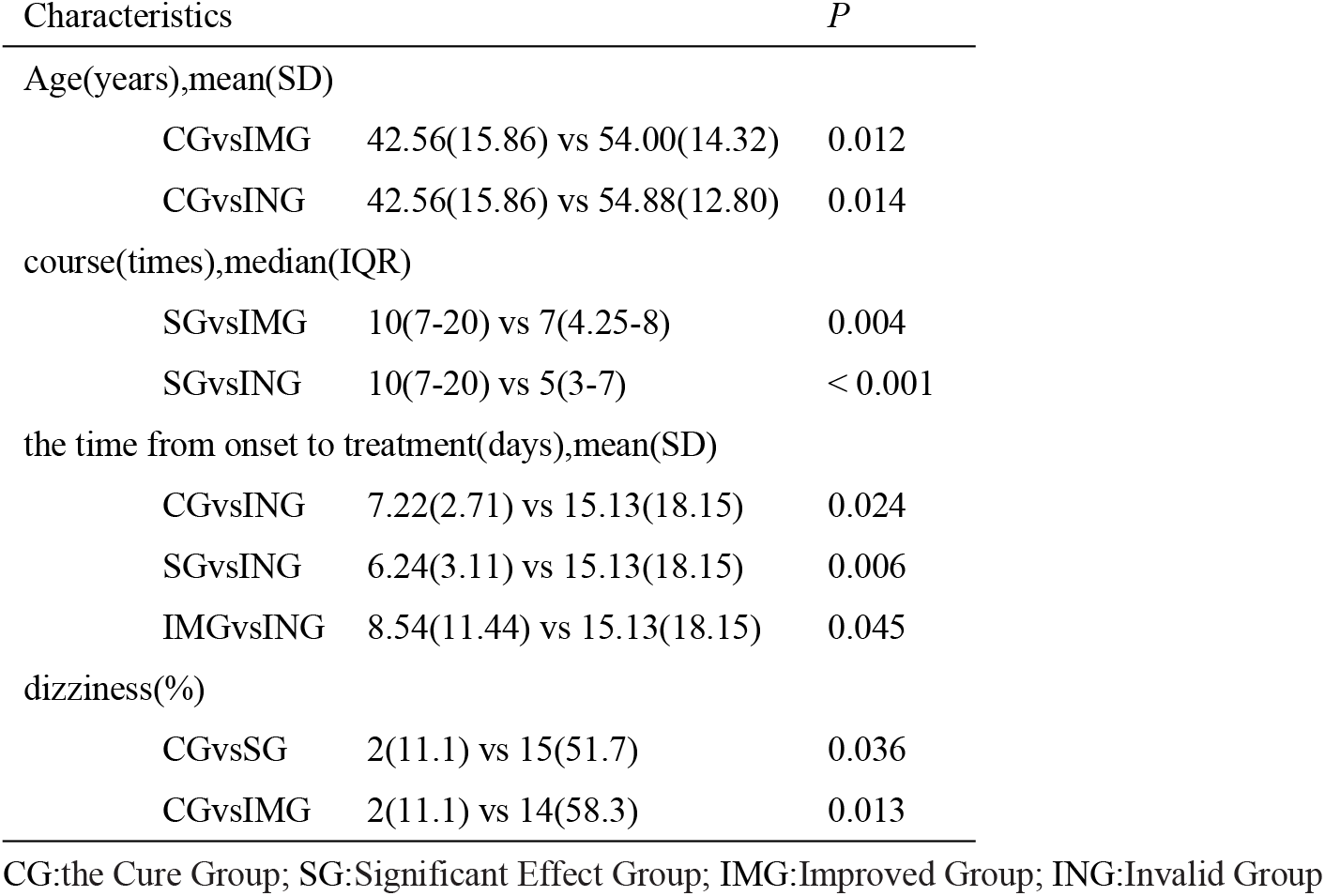
Pairwise multiple comparisons of indicators.

As for the types of audiometric curves, the total effective rate of low frequency descent type was higher than any other types (high frequency descent type, flat descent type, total deafness type)(100% vs 71.4%,81.3%,81.6%, P=0.021)(Table 3).

**Table 3:** Relationship between the types of audiometric curves and prognosis.

| the types | CG(*n* = 18) | SG(*n* = 29) | IMG(*n* = 24) | ING(*n* = 16) | total effective rate(%) |
| --- | --- | --- | --- | --- | --- |
| low frequency descent type | 3 | 3 | 1 | 0 | 100 |
| high frequency descent type | 1 | 4 | 5 | 4 | 71.4 |
| flat descent type | 12 | 9 | 5 | 6 | 81.3 |
| total deafness type | 2 | 13 | 13 | 6 | 81.6 |

*P* = 0.021
CG:the Cure Group; SG:Significant Effect Group; IMG:Improved Group; ING:Invalid Group

In our study, the degree of hearing impairment had no statistical difference with clinical outcome(P=0.073) (Table 4).

**Table 4:** Relationship between the degree of hearing impairment and prognosis.

| the degree | CG( *n*= 18) | SG(*n* = 29) | IMG(*n* = 24) | ING(*n* = 16) |
| --- | --- | --- | --- | --- |
| 30-50 | 7 | 8 | 7 | 8 |
| 51-70 | 9 | 12 | 7 | 1 |
| 71-90 | 2 | 9 | 9 | 6 |
| >90 | 0 | 0 | 1 | 1 |

*P* = 0.073
CG:the Cure Group; SG:Significant Effect Group; IMG:Improved Group; ING:Invalid Group

Age, treatment course, the time from onset to treatment, with dizziness or not, hypertension, the degree of hearing impairment, the types of audiometric curves differed significantly with a P value less than 0.10 were selected as covariates for ordered logistic regression analysis. The results were presented in Table 5. The low frequency descent type and appropriate long treatment course were protective factors to good outcomes(OR = 7.958, 95% CI = 1.148-55.186, P =0.036; OR = 1.089, 95% CI = 1.017-1.167, P =0.015). The elder patients and the longer time from onset to treatment were risk factors to poor outcomes(OR= 0.964, 95% CI =0.934-0.995, P =0.023; OR = 0.919, 95% CI = 0.867-0.975, P =0.005).

**Table 5:** Ordered logistic regression analysis for the prognosis of SHL.

| Variables | OR (95%CI) | P |
| --- | --- | --- |
| the types of audiometric curves |  |  |
| low frequency descent type | 7.958(1.148-55.186) | 0.036* |
| high frequency descent type | 0.630(0.134-2.958) | 0.559 |
| flat descent type | 2.948(0.647-13.435) | 0.162 |
| Age(years) | 0.964(0.934-0.995) | 0.023* |
| course(times) | 1.089(1.017-1.167) | 0.015* |
| the time from onset to treatment(days) | 0.919(0.867-0.975) | 0.005* |

SHL:sudden hearing loss; CG:the Cure Group; SG:Significant Effect Group; IMG:Improved Group; ING:Invalid Group
\*: $P < 0.05$

## Discussion

The pathogenesis and pathological changes of SHL are complicated. Some researchers had confirmed through the development of pathological models of SHL that viruses could induce changes in the stria vascularis and cause inflammatory alterations in nerve cells.^[14]^ Additionally, some scholars believed that SHL was associated with various vascular and coagulation disorders. The function of the inner ear was significantly affected by ischemia, as its blood supply relied on the end arterioles.^[15]^ However, the mechanisms of HBO treatment can effectively address many aspects of SHL’s underlying causes. Firstly, HBO enhances blood supply to the inner ear, increases blood oxygen levels, and alleviates ischemic and hypoxic conditions in this region. ^[16]^Secondly, it boosts the activity of antioxidant enzymes, inhibits the formation of reactive oxygen species, and enhances overall antioxidant capacity, thereby preventing oxidative stress and redox imbalances that can harm cochlear hair cells.^[17]^ Additionally, HBO therapy stimulates stem cells and growth factors, suppresses inflammatory responses, and improves microcirculation.^[18, 19]^ In 2012, the European Undersea and Baromedical Society recognized SHL as an indication for HBO therapy.^[12]^ The latest guidelines from the United States, published in 2019, recommend that HBO treatment combined with corticosteroid therapy be considered a first-line intervention within two weeks of symptom onset. ^[5]^ These guidelines underscored the importance of raising awareness about the potential benefits of HBO therapy for SHL.

In our study, young people were more likely to get a good outcome. Sherlock et al had reported that patients younger than 50 years of age had more significant hearing improvement than patients older than 50 years of age among 78 HBO therapy patients.^[20]^ Aslan et al had found that patients under 50 years old after HBO therapy, hearing level increased significantly.^[21]^ Some studies had shown that patients over the age of 60 had lower recovery rates after treatment. This might be attributed to the fact that the auditory system in the elderly is more degraded, resulting in a weakened capacity for compensatory repair and a reduced sensitivity to treatment.

From our results, we found that SG had a longer treatment course than IMG and ING. But there were no statistical difference between CG and SG, and even the medians of CG(medians 7.5) were less than SG(medians 10). This indicated that, within an appropriate range, a longer duration of treatment yields better outcomes. However, once this range was exceeded, extending the treatment duration did not result in additional benefits. This phenomenon might be linked to severe hearing loss and its irreversible nature. Based on our research, we suggested that a treatment course of 10 to 20 was more favorable.

Capuano et al found that patients treated with HBO therapy within 14 days of onset recovered significantly better than those treated 14 days later.^[22, 23]^ Our findings were similar(Table 2). Therefore, early HBO therapy was particularly important.

Most of SHL patients accompanied by tinnitus and/or dizziness. ^[23]^In our study, tinnitus did not affect prognosis. But patients who suffered dizziness were more likely to lead a poor outcome. The presence of dizziness indicated that the disease had affected the vestibular region, suggesting a more severe condition in patients. Consequently, the occurrence of dizziness could serve as a valuable prognostic indicator.

As for the types of audiometric curves, the total effective rate of low frequency descent type was higher than any other types (P=0.021) reached 100%. But that of high frequency descent type was only 71.4%. The cochlea has a relatively limited blood supply and requires a significant amount of oxygen, making it susceptible to hypoxia. A research by Gupta et al showed, a low birth weight of infants (<2.5 kg) was associated with higher risk of adult-onset hearing loss, because of the poor cochlear development.^[24]^ High-frequency hearing is primarily associated with the basal region of the cochlea, whereas low-frequency hearing is linked to the apical region. Mattox and Simmons^[25]^ showed that the ability to recover the apex was perhaps better than the basal cochlea, the functional metabolic needs of these 2 structural parts might be different, or that the blood supply to distinct parts of the cochlea might be different.^[26]^

Fujimura et al had reported that When the initial hearing loss was more than 80 dB, HBO combination therapy was significantly more effective than drug therapy alone.^[27]^ A similar finding was found by Adjuk et al that when the hearing loss was more than 60 dB, the hearing was significantly improved after HBO therapy.^[28]^ It appeared that more severe hearing loss might lead to better outcomes with HBO. However, some domestic researchers had reached an opposing conclusion. To date, our research had not found a correlation between the degree of hearing loss and clinical outcomes.

Today oxygen was still the only known method of increasing the oxygen level of the inner ear.^[29]^ A large of recent evidences suggested that HBO therapy combined with oral or tympanic corticosteroids could enhance hearing recovery in patients with SHL compared to corticosteroids alone.^[7, 26]^ The results of ordered logistic regression analysis indicated that factors such as the types of audiometric curves, age, HBO treatment course, and the time from onset to treatment were relevant to the outcomes of HBO combination therapy.

Our study has its limitations. Our research is a single‐center study and sample size may not be large enough to represent the general SHL population. We selected an optimal pressure within the therapeutic range of the hyperbaric oxygen chamber, but did not conduct research on varying pressure levels. Further work will be done to verify these findings in more SHL patients.

## Conclusion

In summary, factors such as low frequency descent type‚younger age‚an appropriately longer treatment duration, absence of dizziness and timely initiation of treatment were relevant to a good clinical outcome. A treatment course of 10 to 20 HBO therapy(2ATA, 30 minutes twice) might be more suitable. Therefore, early detection, prompt treatment, appropriate treatment course and individualized strategies for managing SHL were of great significance.

## Data Availability

All relevant data are within the manuscript and its Supporting Information files.

## Acknowledgments

We acknowledge Ningbo Medical Center Li Huili Hospital for supporting our research.

## Institutional Review Board Statement

As a retrospective study, this research was conducted in accordance with the Declaration of Helsinki, and approved by the Ethics Committee of Ningbo Medical Center Lihuili Hospital (Approval No.: LHHL-EC-2024-Research-520) with a waiver of patient informed consent.

